# Outcomes of Advanced Malignant Melanoma in a South African Public Health Sector Cohort: The Urgent Need for Effective Therapy

**DOI:** 10.1101/2020.07.16.20155366

**Authors:** Thulo Molefi, Cert Med

**Affiliations:** Department of Medical Oncology, Faculty of Health Sciences, University of Pretoria, Pretoria, South Africa

## Abstract

**Background:** Immunotherapy and molecularly targeted therapy have revolutionised the treatment of malignant melanoma, however for South Africa’s public health sector patient population, these treatment modalities are far out of reach and chemotherapy remains the only treatment option.

**Aim:** To evaluate the outcomes of advanced melanoma and determine the need for therapies other than conventional chemotherapy in South Africa’s public health sector.

**Setting:** The Department of Medical Oncology, Steve Biko Academic hospital (SBAH), Pretoria, South Africa.

**Methods:** Files of patients with advanced malignant melanoma managed at SBAH, from 01 January 2009 to 31 December 2019 were retrospectively reviewed.

**Results:** One hundred files meeting the inclusion criteria were analysed, 24 with regional (stage III) and 76 with metastatic (stage IV) disease. 23 (96%) patients with regional disease didn’t receive adjuvant therapy and had a median time to progression (mTTP) of 12 months (95%CI; 8.9-15.0). Within the metastatic melanoma cohort, 34 (79.1%) patients received chemotherapy and had a median overall survival (mOS) of 5 months (95% CI; 4.3-5.6), while patients that didn’t receive chemotherapy had a mOS of 2 months (95% CI; 0.8-3.1) (p=0.213).

**Conclusion:** These results reaffirm the impotent effects of chemotherapy in treating malignant melanoma and it is imperative that South Africa’s public health sector expands its armamentarium against this lethal disease.

## Introduction

South Africa (S.A) is a country with high solar ultraviolet radiation levels,^[1]^ which is a known risk factor for malignant melanoma. Melanin is a protective factor against developing skin malignancies and in S.A, with its racially diverse population predominated by black Africans,^[2]^ it’s not astonishing that there’s a racial disparity in the incidence of melanoma.^[3]^While the incidence is lower in black South Africans, this population is more likely to have an aggressive disease process and often present at an advanced stage.^[4]^

The South African health system is divided into public and private sectors, with the countries indigent citizens being treated in the public sector.^[5]^ In the public sector drugs available for the management of a wide range of diseases are listed on an essential medicines list (EML), however malignant melanoma and it’s treatment is not included in the countries national EML.[13]

Immunotherapy and molecularly targeted therapy have revolutionised the treatment of malignant melanoma, by lowering the risk of recurrence in regional (stage III) disease^[6,7]^ and significantly prolonging survival in patients with metastases (stage IV),^[8-12]^ however for South Africa’s public health sector patients’, these treatment modalities are far out of reach and chemotherapy with its known low and short-lived responses,^[14,15,16]^ remains the only treatment option.

Previously published South African studies into melanoma mainly focused on epidemiological data^[1,4,17-20]^ or discussed surgical management of localised disease,^[21]^ with no studies looking at the treatment outcomes, particularly that of advanced disease (stage III/IV). Steve Biko Academic hospital’s (SBAH) medical oncology department, as a tertiary referral hospital department and one of a few cancer units in S.A’s public health sector, is responsible for the management of many of these patients.

This study evaluated the outcomes of advanced melanoma in patient’s managed at SBAH from 01 January 2009 to 31 December 2019, in order to determine the need for therapies other than conventional chemotherapy in S.A’s public health sector.

## Methods

A retrospective review of patient files with advanced (stage III/IV), histologically confirmed malignant melanoma managed at SBAH from 01 January 2009 to 31 December 2019, was conducted to evaluate the outcomes. Patients were identified using the department’s referral register for the study period and once identified, the files were manually searched for at records filing. Patient file numbers documented on the departments referral register but not found at records, missing histology reports, localised disease and patients treated on a clinical trial or received prior therapy at another institution were excluded.

Demographic, clinical, radiological and treatment response data was collected and entered on an excel spreadsheet prior to being entered into IBM SPSS version 26 for statistical analysis. Time to progression (TTP) was defined as the time from first visit until the detection of recurrence or metastases and overall survival (OS) was defined as the time from initial visit until death.

Ethical approval to conduct the study was obtained from the Faculty of Health Sciences Research Ethics Committee, University of Pretoria (Ethics reference No. 158/2020).

## Results

A total of 313 patients were recorded on the department’s patient register as having been seen for malignant melanoma from 01 December 2009 until 31 December 2019. 85 files were not found, histology was missing from 2 files, 63 files had incomplete staging, 58 had localised disease and 105 had advanced melanoma. 5 patients from the stage IV cohort had received chemotherapy prior to their referral to SBAH, leaving 76 metastatic and 24 regional melanoma files meeting the inclusion criteria (figure 1).

**Figure 1:**
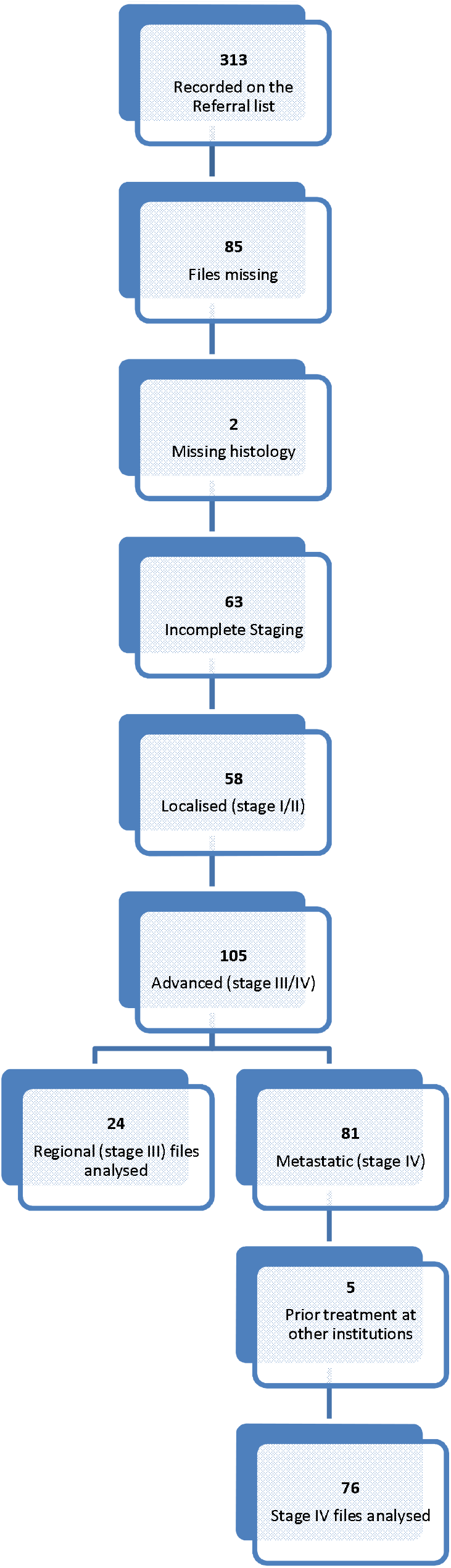
Study Outline

The regional melanoma cohort had a median age of 54 years (range 15-82), consisted of 17 (71%) female and 7 (29%) male patients, 18 (75%) of whom were caucasian and 6 (25%) black Africans. 18 (75%) of the patients had a wide local excision (WLE) of their primary lesion together with a lymph node dissection (LND) of the regional lymph nodes, while 3 (12.5%) presented with lymphadenopathy and had LND only and the remaining 3 (12.5%) only had a WLE.

Majority of the patients (23 (96%)) didn’t receive adjuvant therapy after the surgical management of their disease with 1 (4%) patient, receiving adjuvant radiotherapy (RT) to her axilla. This patient developed lymph node (LN) recurrence in the radiated axilla 17 months later, based on positron emission tomography/computed tomography (PET/CT) findings, however this biopsied for histological confirmation as she had significant lymphedema post her initial LND and adjuvant RT.

During follow up post surgical management, thirteen (54.1%) patients were lost to follow up, 9 (37.5%) developed metastases at a median of 12 months (95%CI; 8.9-15.0), while 2 (8.3%) patients were still in remission at the time of analysis (figure 2).

**Figure 2:**
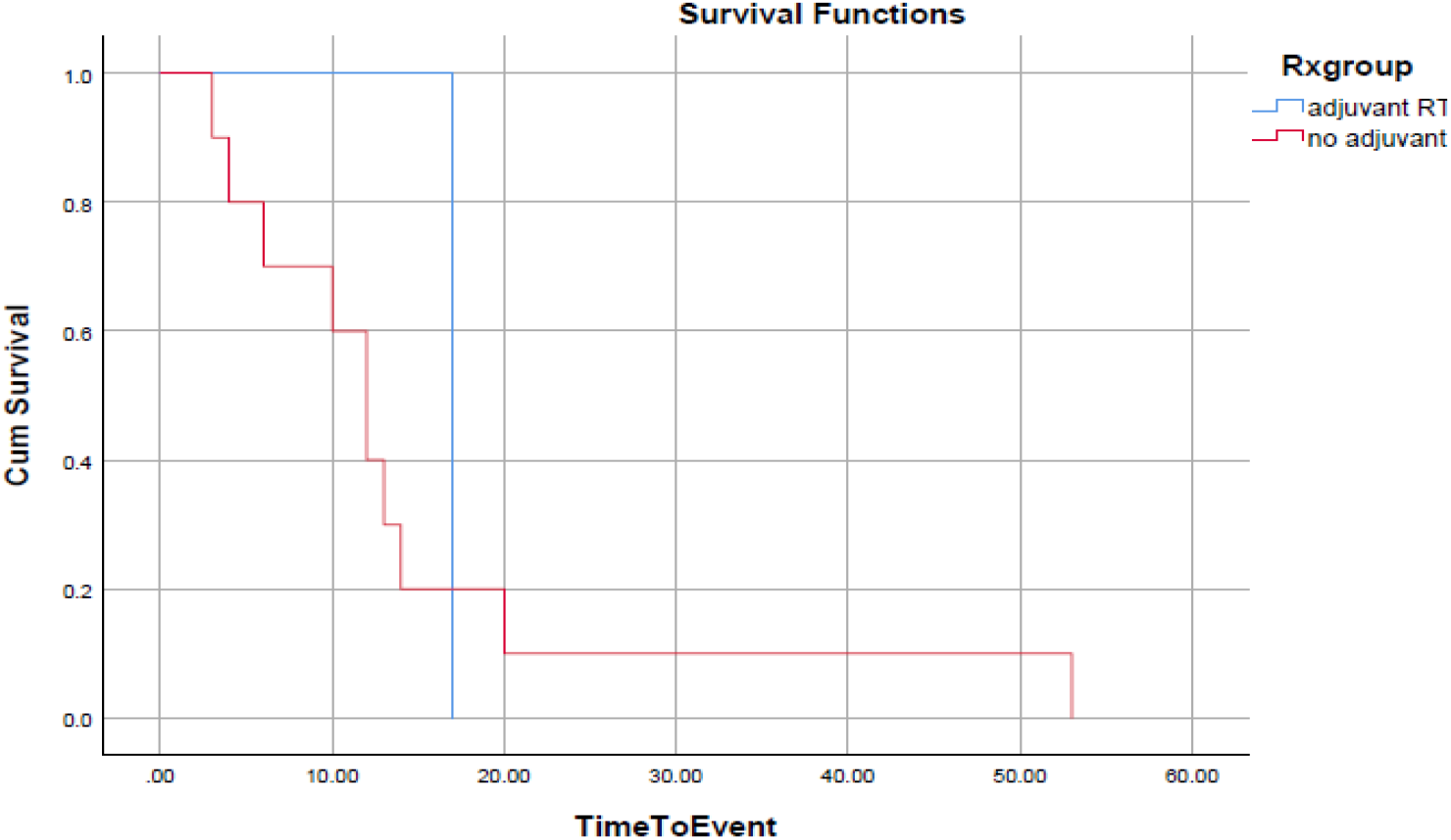
Stage III time to progression (TTP)

In the de-novo metastatic cohort, patients had a median age of 53.6 years (range 19-88), comprising of 44 (57.8%) males and 32 (42.2%) females, 32 and 44 of whom were of the black and white races, respectively. Forty three (56.6%) received treatment, 14 (18.4%) patients were not fit for therapy and had supportive care, while 6 (7.9%) refused chemotherapy and 13 (17.1%) were lost to follow up shortly after their initial consultations.

The patient population that received therapy, 9 (20.9%) had brain metastases and were referred for whole brain irradiation (WBI) and 3 of them succumb to their disease with a median overall survival (mOS) of 1 month (figure 3), while the remainder were lost to follow up.

**Figure 3:**
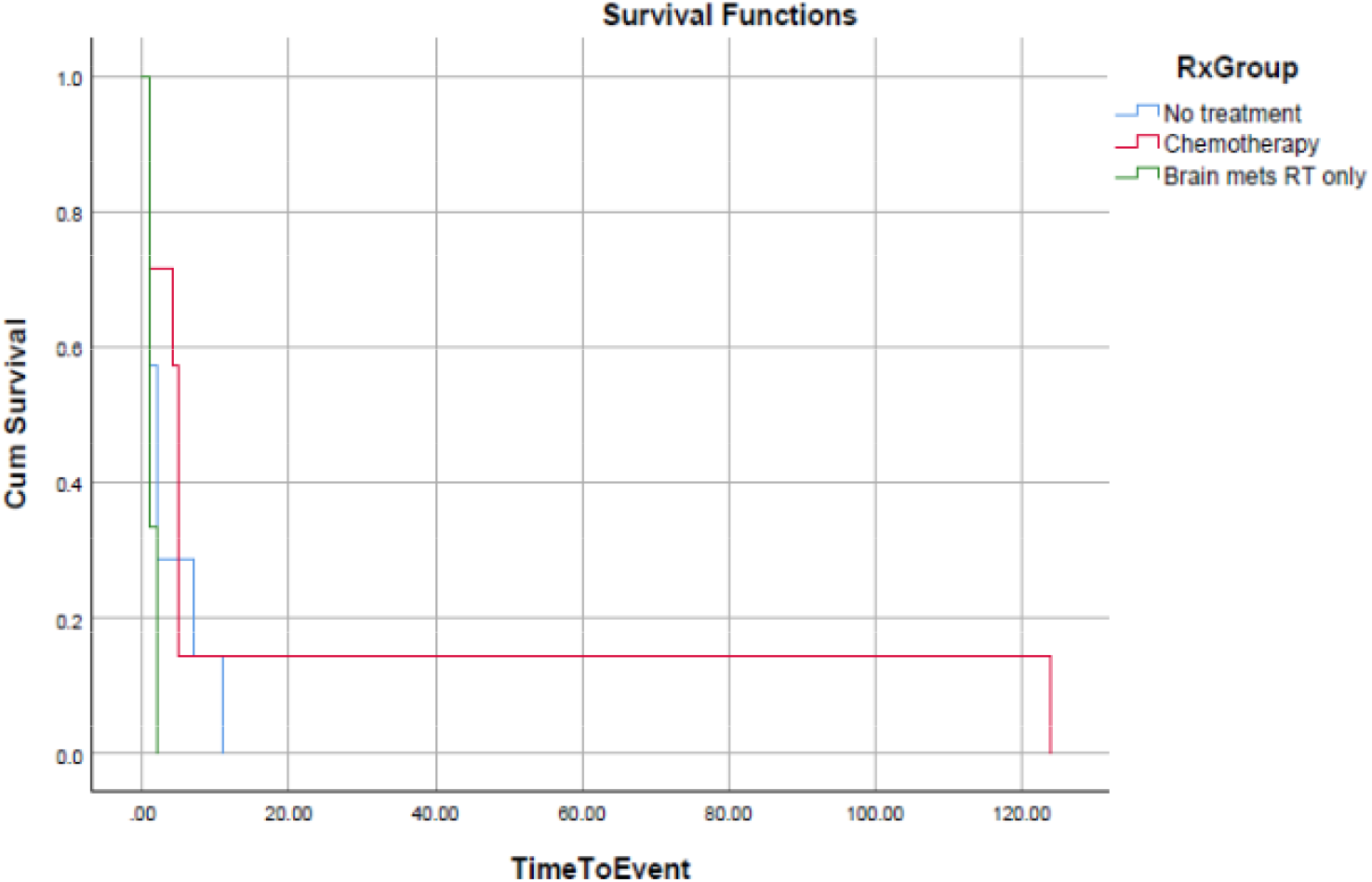
Stage IV overall survival (OS)

Thirty four (79.1%) of patients with non-brain metastases received chemotherapy and had a median overall survival (mOS) of 5 months (95% CI; 4.3-5.6)(figure 3), 28 received dacarbazine (DTIC) every three weeks and 6 received a carboplatin-paclitaxel (C-P) doublet every three weeks. Ten patients were lost to follow up and three patients demised while on initial chemotherapy. Nineteen patients had disease progression while on chemotherapy (3 C-P and 16 DTIC), 3 of whom died post progression and the remainder were lost to follow up.

One patient was still on treatment at the time of analysis and another patient was still in remission 10 years after her initial visit. The patient still in remission was first seen in April of 2009, after her diagnosis was confirmed on a liver biopsy a month earlier with no other detectable disease clinically and on a PET/CT. She received 26 cycles of DTIC with no documented adverse effects and in 2012, when the DTIC was stopped; she still hadn’t progressed and was referred for selective internal radiation therapy (SIRT). In 2013 after SIRT, a PET/CT still detected isolated liver lesions followed by a biopsy showing fibrosis. The liver still had flourodeoxyglucose (FDG) lesions on a PET/CT in 2017 which were observed and she was still alive and well at her last consultation in 2019.

The cohort that didn’t receive chemotherapy had a mOS of 2 months (95% CI; 0.8-3.1), which is statistically indistinguishable from the cohort that received chemotherapy (p=0.213) (figure 3). The 14 patients offered supportive care, 9 (64.2%) were lost to follow up and 5 (35.8%) demised, while the 6 patients who declined chemotherapy, 1 patient succumbed to his disease 11 months after his initial visit, 4 were lost to follow up with the remaining patient, first seen in December 2019, with multi-organ metastases and was still alive at the time of data analysis.

## Conclusion

Internationally and in South Africa’s private health sector, the current treatment landscape for malignant melanoma has relegated chemotherapy to the ignored outskirts of therapeutic options,^[22,23]^ however the lack of options in South Africa’s public health sector^[13]^ still leads to its continued use.

Drugs used as the standard of care in the adjuvant setting have delayed progression, with 3- year relapse free survival rates of up to 58%,^[6,7]^ and those used in patients with metastatic melanoma have improved overall survival over 60 months,^[8,9,10,11,12]^ while S.A’s public health sector population do not have access to these therapies and relapse in a median of 12 months or perish from metastatic melanoma at a median of 5 months, because of ineffective therapy.

This study’s results reaffirm the impotent effects of chemotherapy in treating malignant melanoma and it is imperative that South Africa’s public health sector expands its armamentarium against this lethal disease.

## Data Availability

Demographic, clinical, radiological and treatment response data was collected and entered on an excel spreadsheet prior to being entered into IBM SPSS version 26 for statistical analysis.

## Disclaimer

The views expressed in this article are those of the author and are not on behalf of the institution where the study was performed.

## Supplementary: Figures

